# Perceptions of persons deprived of liberty regarding tuberculosis vaccine research

**DOI:** 10.1101/2025.07.04.25330885

**Authors:** Mariana Cristina Campos Falleiros Pires, Yiran E Liu, Everton Ferreira Lemos, Liliane Ferreira da Silva, Mariana Garcia Croda, Monica Magalhães, Dhélio Batista Pereira, Mariana Pinheiro Alves Vasconcelos, Rosilene Ruffato, Solana Monteiro Batista, Giselle Lima de Freitas, Marcelo Cordeiro dos Santos, Giane Zupellari Santos-Melo, Jair dos Santos Pinheiro, Lara Bezerra de Oliveira Assis, Lia Gonçalves Possuelo, Tiago Antonio Heringer, Daiane Kist Back, Pauline Schwartzbold, Jason R Andrews, Crhistinne Cavalheiro Maymone Gonçalves, Julio Croda

## Abstract

**Background:** Several tuberculosis (TB) vaccine candidates are currently advancing to late stage clinical trials. Prisons in low- and middle-income countries harbor some of the highest rates of TB in the world, making persons deprived of liberty (PDL) an important population to prioritize for the introduction of effective vaccines. However, their inclusion in clinical trials raises significant ethical concerns due to a history of exploitation and mistreatment within medical research. To date, PDL’s own perspectives on participating in vaccine research have been largely overlooked. This study aimed to understand the perceptions of PDL regarding TB, vaccines, and their potential participation in clinical trials of new TB vaccines.

**Methods:** This multicenter qualitative study employed focus group (FG) discussions in seven state prisons across four of the five regions of Brazil, involving 91 incarcerated individuals (64 men and 27 women). Participants were selected through convenience sampling and interviewed between February and August 2024. The FGs followed a standardized structure, exploring participants’ perceptions regarding health in prisons, TB, vaccines in general, new TB vaccines, and their potential participation in research. The discussions were transcribed verbatim and analyzed using thematic content analysis.

**Results:** Participants reported encountering difficulties in accessing healthcare services within the prison system. They also shared personal or indirect experiences with TB, as well concerns about their family members being at risk for tuberculosis exposure. While participants generally held positive perceptions about vaccines and vaccine trials, they emphasized the need for clear and transparent information, respect for individual autonomy, and assurances of accountability from researchers as conditions of their willingness to participate in future TB vaccine trials.

**Conclusions:** PDL perceptions regarding participation in clinical trials for new TB vaccines are significantly influenced by their prior experiences with the prison health system and their level of trust in research institutions. To ethically and effectively include PDL in future research, it is crucial to prioritize respect for participant autonomy and transparent communication about the risks and potential benefits involved.

## Introduction

Tuberculosis (TB) remains a significant public health concern worldwide, causing an estimated 1.25 million deaths and 10.8 million new cases in 2023(1). It disproportionately impacts persons deprived of liberty (PDL), particularly in Latin America, where prison populations have grown by nearly 300% in the past three decades(1,2). The conditions of imprisonment, such as overcrowding and malnutrition, increase the risk of PDL acquiring TB infection and disease. In South America, the risk of TB is 26 times higher in prisons compared to the general population(3). Furthermore, TB in prisons can spill over into communities through staff, visitors, and individuals released from incarceration, who have an elevated risk of TB for up to seven years post-release (4,5). A recent mathematical modeling study found that incarceration is a significant driver of population-level TB in Latin America, accounting for an estimated 27% of new cases, more than any other risk factor(6). In addition, a genomic epidemiologic study conducted in Brazil found that up to 50% of community-based TB cases may originate from transmission within prison settings (7). Ending the TB epidemic will require comprehensive efforts to address TB in prisons.

To date, interventions to reduce TB in high-burden prisons have been lacking. Structural reforms such as reducing incarceration rates and overcrowding are critical for mitigating transmission(6) but present political challenges often beyond the purview of national TB programs. Systematic screening for TB in prisons, now strongly recommended by the World Health Organization (WHO), has demonstrated high yield in empirical studies in Latin American prisons (8). However, serial rounds of annual mass screening in three Brazilian prisons did not significantly reduce TB prevalence, suggesting the need for more frequent screening and complementary measures(9). Tuberculosis preventive therapy (TPT) presents as a promising strategy(10), but evidence of its effectiveness in high-transmission prisons remains limited(11).

To meaningfully reduce TB in prisons, new tools, such as effective vaccines, may be needed to complement existing measures. The only licensed TB vaccine, BCG, was discovered a century ago and is inadequate in preventing TB in adolescents and adults(12). The current TB vaccine development pipeline has several promising candidates, including M72/AS01, MTBVAC, and VPM1002, which have advanced to phase 3 trials after demonstrating favorable safety profiles and preliminary evidence of efficacy(13–15). However, TB vaccine trials face significant challenges. The relatively low incidence of TB, even in high-burden community settings, necessitates the recruitment of thousands of participants and prolonged follow-up periods to achieve adequate statistical power. This results in slow and costly trial processes that hinder rapid advancements in vaccine development.

PDL may significantly benefit from effective TB vaccines and are recognized as one of many high-risk, target populations to be prioritized for rapid access to new TB vaccines once licensed(16). However, in contrast to other priority populations—like persons living with HIV, persons with diabetes, and persons experiencing malnutrition—PDL have been systematically excluded from participation in TB vaccine trials. Including PDL in vaccine trials presents serious ethical challenges. Historically, PDL have been subjected to exploitative, non-consensual experimentation(17–19). Barriers to privacy, autonomy, and access to healthcare in prison further complicate ethical trial implementation. During the COVID-19 pandemic, as PDL experienced elevated rates of COVID-19 infection and mortality, debates emerged around the risks and benefits of including PDL in COVID-19 vaccine trials, and the necessary conditions to make ethical participation possible(20–23). Despite agreement on the critical importance of engaging PDL in these debates, the perspectives of PDL toward participation in vaccine trials, or scientific research more broadly, remain poorly understood, especially outside of high-income countries. This omission is particularly striking given that tuberculosis remains the leading cause of death from an infectious disease among adults worldwide (24,25), and that recent perspectives have underscored both the injustice of global TB disparities (25) and the ethical imperative to evaluate ultra-short TB prevention regimens with careful scientific and ethical scrutiny(26).

In Brazil, where the worsening TB epidemic is increasingly driven by incarceration, understanding the experiences and perspectives of PDL will be crucial to inform effective, ethical, and patient-centered strategies to accelerate TB elimination. In this study, we conducted qualitative focus group (FG) discussions in seven male and female prisons in Brazil to understand perceptions toward TB, TB research, and participation in TB vaccine trials.

## Materials and Methods

### Study Setting and Population

In this multicenter qualitative study, we conducted FG discussions in five male prisons and two female prisons in Brazil between February and August 2024. The participating prisons were located in Campo Grande (state of Mato Grosso do Sul), Manaus (Amazonas), Montenegro (Rio Grande do Sul), Porto Velho (Rondônia), and Ribeirão das Neves (Minas Gerais) (Fig.1).

**Figure 1.**
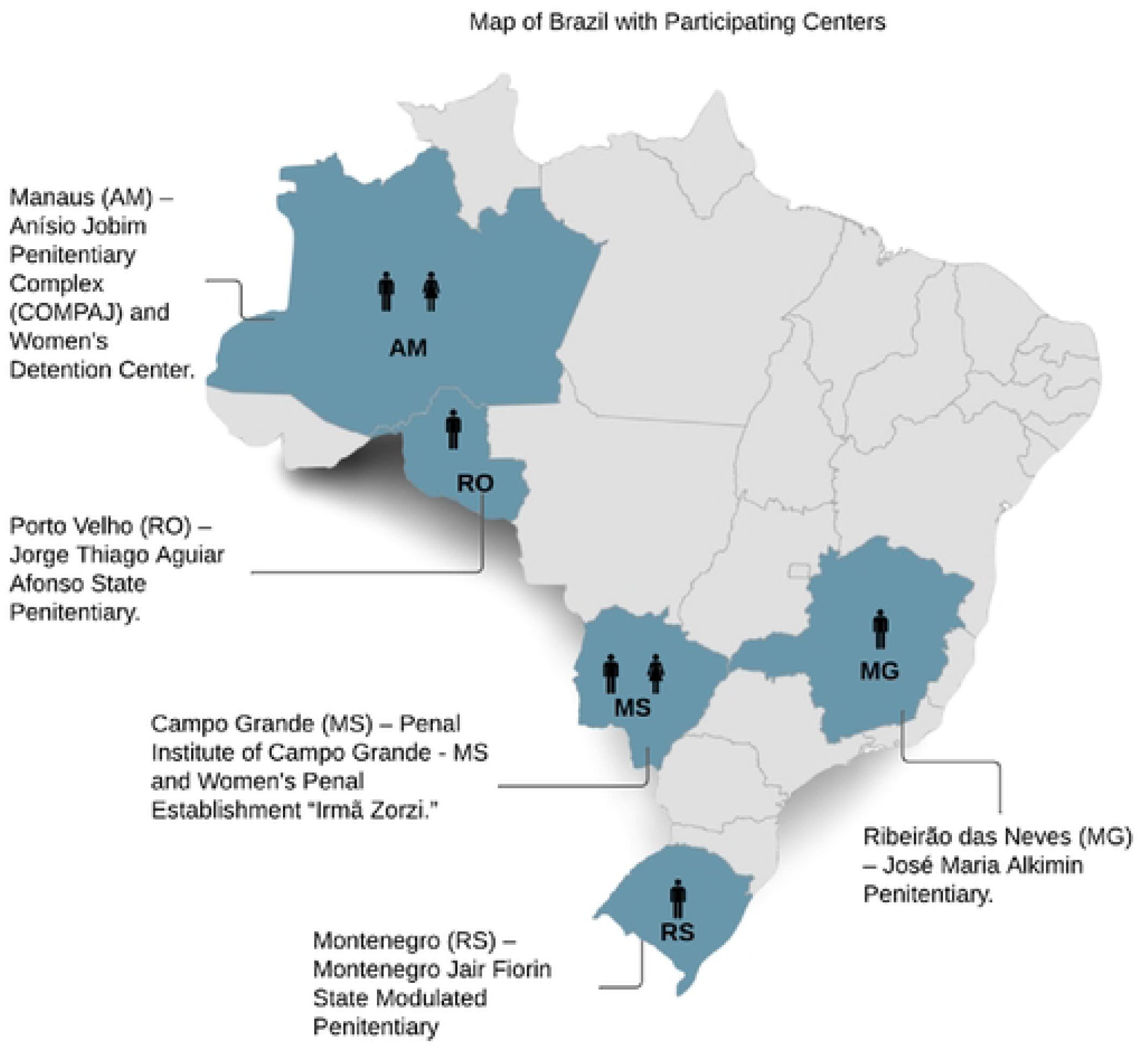
Map of Brazil with study centers

Participants (N=91) were recruited through convenience sampling. Among those who volunteered to participate in the study, participants were selected to ensure male and female inclusion, and representation from diverse cells and pavilions. Additionally, the following inclusion criteria were applied: age 18 or older, literate, have been incarcerated for at least 12 months cumulatively (continuous or non-continuous), and provided voluntary informed consent.

### Study Procedures

To develop the FG question script (Supplementary Appendix 1, Focus Group Questions), we adapted the Vaccine Hesitation Matrix (VHM), developed by the WHO Strategic Advisory Group of Experts on Immunization(27). The script was divided into three main dimensions, which were further subdivided into two categories each:

1. perceptions of health in the prison (prison health services, experiences with TB)
2. perceptions of vaccines (vaccines in general, new TB vaccines)
3. perceptions of participation in research (clinical trials for a new TB vaccine, PDL autonomy).

The full list of FG questions is provided in the Appendix.

Each focus group was conducted by two facilitators and two observers, in private settings within the prisons. The facilitators were experienced in working in prison environments, qualitative health research, including focus group discussions. Observers took field notes during each focus group.

All focus groups followed a standardized structure. Participants were first informed of the study’s objectives and requirements. The facilitators read the Informed consent form aloud, addressing questions from potential participants. Those who decided to participate signed the consent form.

Next, study participants were interviewed individually in private areas using a structured questionnaire designed to collect clinical and epidemiological data. Variables investigated included: sex, age, race/ethnicity, marital status, education level, household income, city/state of residence before incarceration, comorbidities, risk factors, duration of incarceration, transfers between prisons, visitation status, history of contact with a TB patient in the same cell, current TB treatment status, past TB treatment, and use of medications.

The focus group discussion then began, with audio recording. Each group consisted of 12 to 15 individuals and lasted approximately 90 minutes on average. To protect confidentiality, participants were assigned random numbers and instructed to identify themselves by their assigned number before speaking. Any names mentioned during the sessions were removed from the transcripts.

### Data processing and analysis

Questionnaire data were stored in a secure REDCap® server with controlled access. Audio recordings of FG discussions were stored in a secure password-protected folder accessible only to the research team. Transcriptions of FG discussions were performed verbatim by two researchers, maintaining authenticity and literal accuracy. The data were analyzed using the thematic content analysis method proposed by Minayo (2005)(28), following three steps:

1. Pre-analysis: Thorough and detailed reading of the transcripts.
2. Material exploration: Extracting and coding key segments of speech, and grouping them into thematic categories based on recurrence, relevance, and uniqueness.
3. Result processing: Synthesizing and interpreting themes using updated theories and scientific literature to draw robust and relevant insights.

The transcriptions were reviewed and coded by two coders and grouped by thematic areas throughout the analysis, resulting in the emergence of specific dimensions, categories, and subcategories. Triangulation was achieved by holding interactive discussions with facilitators and observers and sharing preliminary results in research team meetings to validate the coding. MaxQDA24®(29) software was used to organize and highlight the coded texts.

### Ethical Approvals

The study received approvals from the Ethics Committee for Research (CEP) of Federal University of Mato Grosso do Sul – UFMS (Approval No. 6.612.127, CAAE: 75865823.7.1001.0021, January 12, 2024), the Center for Research in Tropical Medicine of Rondônia – CEPEM (Approval No. 6.633.785, CAAE: 75865823.7.2001.0011, February 3, 2024), the School of Nursing at the Federal University of Minas Gerais – UFMG (Approval No. 6.708.897, CAAE: 75865823.7.2002.5149, March 18, 2024), the University of Santa Cruz do Sul – UNISC (Approval No. 6.711.854, CAAE: 75865823.7.2005.5343, March 19, 2024), and the Tropical Medicine Foundation Doctor Heitor Vieira Dourado – FMT-HVD (Approval No. 6.745.992, CAAE: 75865823.7.2004.0005, April 5, 2024), as well as consent from the State Departments of Penitentiary Administration.

## Results

We conducted a total of seven focus groups with 91 PDL, of which 64 (70%) were men and 27 (30%) women. Participants had an average age of 35 years and many had low education levels (41% with no high school) (Table 1). Half of participants identified as mixed race (50%), followed by white (25%) and black (20%). Regarding duration of incarceration, 41% had been imprisoned for up to 2 years, 18% for 3 to 4 years, 24% for 5 to 9 years and 18% for 10 years or more. The majority of participants (63%) had a history of prior incarceration, 66% had been transferred between prisons, and 67% reported receiving visitors. Among participants, 20% had a prior TB diagnosis, 3% were currently undergoing TB treatment, and 60% reported having had contact with cellmates who had TB. In the prison units where focus groups were conducted, the reported incidence of TB varied greatly, ranging from 685 to 4,533 per 100,000 person-years.

**Table 1:** Clinical and epidemiological characteristics of focus group participants (N=91)

| Characteristic | n (%) |
| --- | --- |
| **Sex** |  |
| Female | 27 (30) |
| Male | 64 (70) |
| **Age group** |  |
| 20-39 years | 62 (68) |
| 40-59 years | 27 (30) |
| 60+ years | 2 (2) |
| **Race** |  |
| Asian | 4 (4) |
| White | 23 (25) |
| Indigenous | 1 (1) |
| Mixed | 45 (50) |
| Black | 18 (20) |
| **Education** |  |
| Incomplete elementary education | 24 (26) |
| Complete elementary education | 13 (14) |
| Incomplete high school | 22 (24) |
| Complete high school | 16 (18) |
| Incomplete higher education | 10 (11) |
| Complete higher education | 2 (2) |
| Postgraduate | 1 (1) |
| Did not respond | 3 (3) |
| **Length of incarceration** |  |
| Up to 2 years | 37 (41) |
| 3-4 years | 16 (18) |
| 5-9 years | 22 (24) |
| 10 years or more | 16 (18) |
| **Transfers between prisons** |  |
| Yes | 60 (66) |
| **Previous incarceration** |  |
| Yes | 57 (63) |
| **Receive visits** |  |
| Yes | 60 (67) |
| **TB history** |  |
| Yes | 18 (20) |
| **Contact with a cellmate with TB** |  |
| Yes | 55 (60) |
| **Undergoing TB treatment** |  |
| Yes | 3 (3) |

Participating PDL described their perceptions regarding the health system in prisons, vaccines in general, a potential new TB vaccine and participation in clinical trials. Representative quotes are presented in Table 2, organized by dimensions, categories, and subcategories that emerged in the analysis.

**Table 2:**
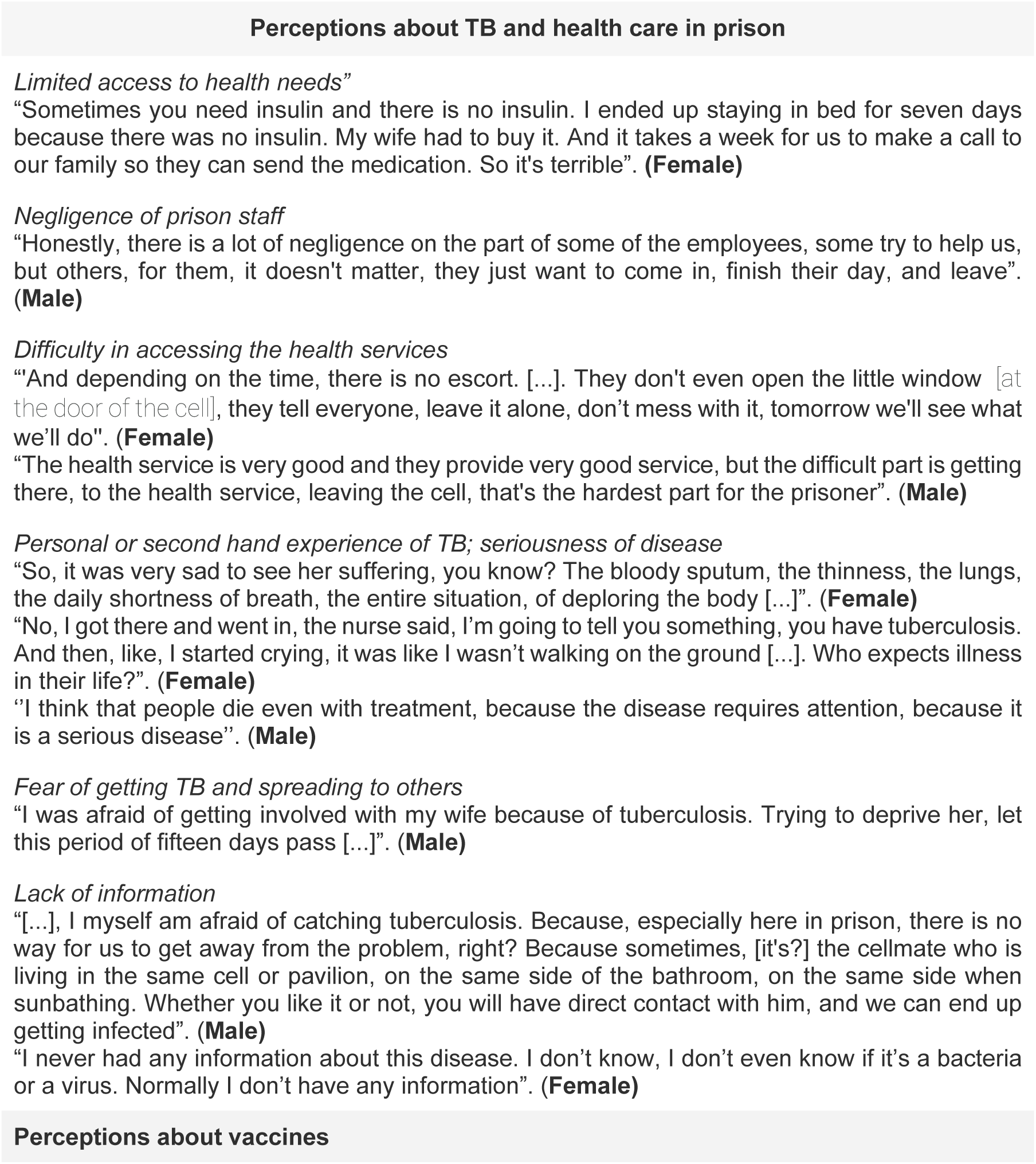
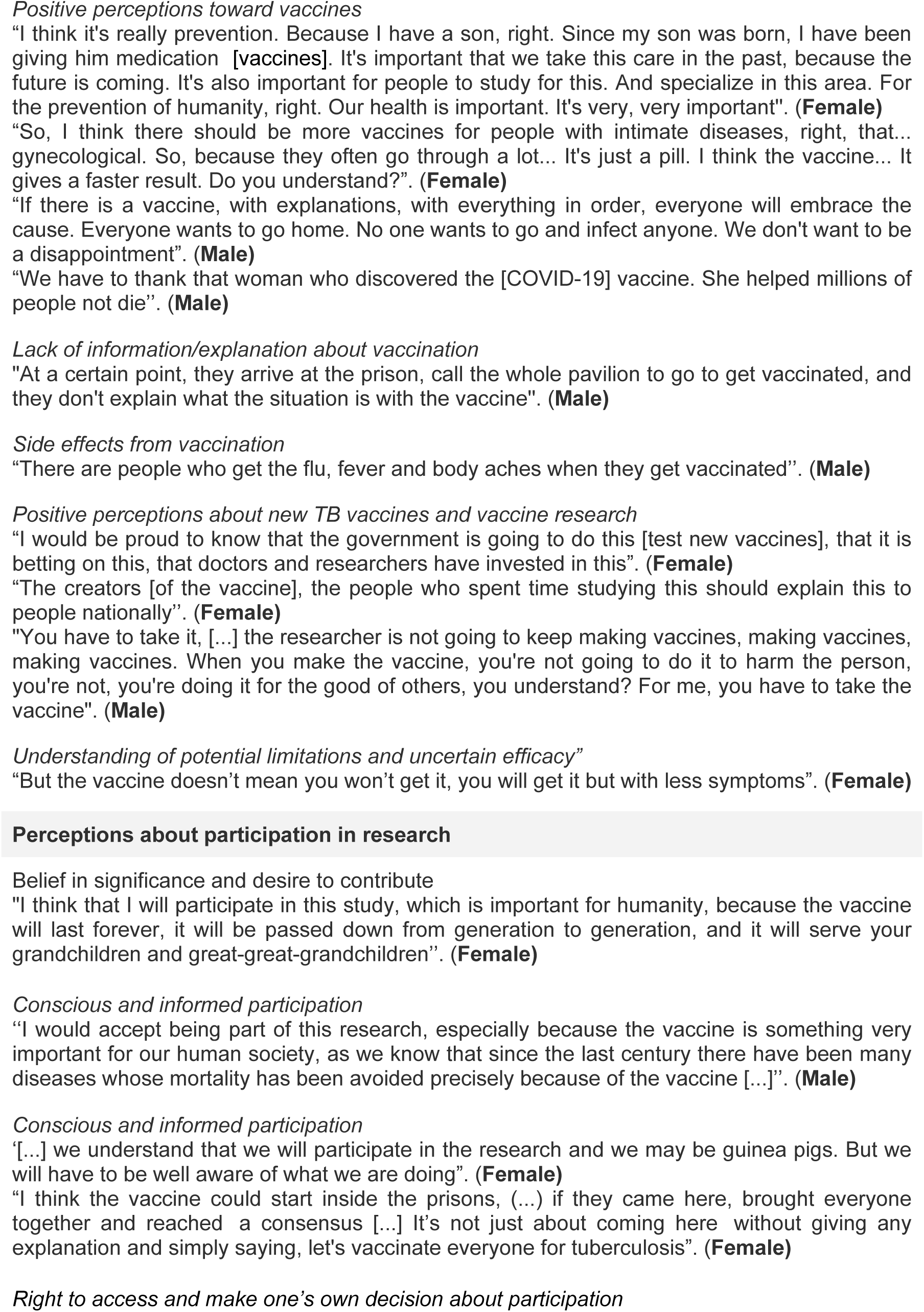
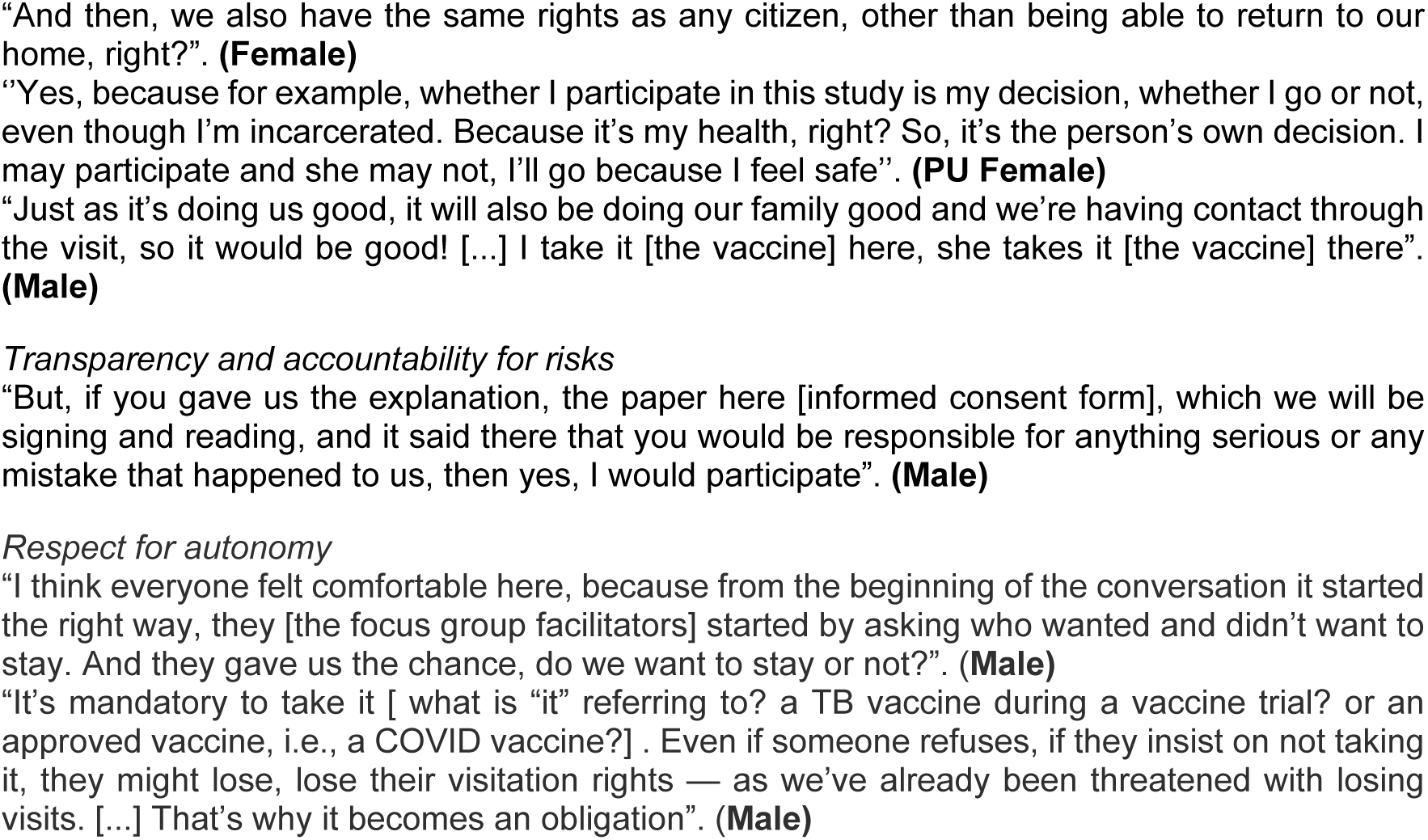
Perceptions of participants regarding health care in prisons, TB, vaccines, and participation in research.

### Perceptions about health in prison

#### Access to health care in prison

Participants described many challenges in accessing health care in prison (Table 2). Access to essential medicines was limited, as one participant described: “Sometimes you need insulin and there is no insulin. I ended up staying in bed for seven days because there was no insulin. My wife had to buy it. And it takes a week for us to make a call to our family so they can send the medication.”

Participants also reported significant difficulties in accessing health services, in part due to factors external to the health team. In particular, participants cited logistical and structural barriers such as clinic hours of operation and the lack of guard escorts to the health unit. As one participant explained, “The health service is very good…but the difficult part is getting there…leaving the cell, that’s the hardest part for the prisoner.” Access to health services could be further hindered by the attitudes and attentiveness (or lack thereof) of prison staff. As one participant put it, “Honestly, there is a lot of negligence on the part of some of the employees, some try to help us, but others, for them, it doesn’t matter, they just want to come in, finish their day, and leave.”

#### Perceptions and experiences of TB

Participants described TB as a serious, potentially fatal disease causing various physical symptoms and requiring rigorous treatment. Accordingly, participants frequently cited fears of developing TB and the pervasiveness of the disease in prison. One participant described the perceived inevitability of exposure as follows: “Here in prison, there is no way for us to get rid of the problem, right? Whether you like it or not, you will have direct contact with [a person with TB], and we can end up getting infected.”

Participants who had personal or secondhand experience of TB reported feelings of dismay upon diagnosis and distress from witnessing the suffering of others with TB. One participant reflected on the moment she was told she had TB: “I started crying…it was like I wasn’t walking on the ground.” Another participant with previous TB expressed concern about transmitting TB to a family member during visits.

#### Lack of information

While participants had a basic understanding of TB as an infectious disease, many reported the difficulty of obtaining accurate information about TB. They highlighted the lack of access to reliable information sources and the absence of educational activities in prison. Some expressed uncertainty about the causative agent of TB, routes of transmission, and effective preventive measures.

### Perceptions of vaccines

#### Positive sentiments toward vaccines

Vaccines were generally viewed favorably by participants, with wide recognition of the importance of vaccination for protecting the health of oneself and one’s family. Scientific advances in immunization, especially for vaccines against COVID-19, were lauded by participants as historic, lifesaving milestones. Some participants also called for the development of vaccines against other diseases, expressing a preference for prevention over treatment.

Regarding the development of new vaccines against TB, participants applauded the efforts of researchers and governments to seek solutions to control the disease, viewing it as a social good for humanity. As one participant stated, “I would be proud to know that the government is going to do this…that doctors and researchers have invested in this.” Another participant expressed confidence in the benevolent motivations of those developing the vaccine: “When you make the vaccine, you’re not going to do it to harm the person…you’re doing it for the good of others.”

#### Perceptions about side effects and efficacy

Participants expressed concerns about possible side effects from vaccination, citing first- or second-hand experience of flu-like symptoms, fever, and body aches. They also reported unease around potential unknown adverse reactions and the lack of follow-up after vaccination. Some participants described the potential limitations of vaccines, recognizing that vaccines may not offer full protection but could reduce the severity of disease.

#### Negative experiences with vaccination in prison

Some participants expressed dissatisfaction or discomfort with prior vaccination efforts within the prison. For instance, one participant recalled an experience of being called to get vaccinated without clear information or understanding: “They arrive at the prison, call the whole pavilion to go to get vaccinated, and they don’t explain what the situation is with the vaccine.”

### Perceptions about participation in research

#### Perceptions toward participation in clinical trials

Many participants expressed a desire to participate in clinical trials for a new TB vaccine, describing it as an opportunity to contribute to an effort they viewed as significant for society. One participant stated, “I think that I will participate in this study, which is important for humanity, because the vaccine will last forever, it will be passed down from generation to generation, and it will serve your grandchildren and great-great-grandchildren.”

Some participants associated participation in clinical trials with the idea of being “guinea pigs” for a vaccine that could turn out to be ineffective. Yet, many expressed a willingness to participate nonetheless, as long as it was with intention and awareness of potential risks and benefits: “We understand that we will participate in the research and we may be guinea pigs. But we will have to be well aware of what we are doing.” Participants gave a variety of reasons for their decision to participate, ranging from a desire for personal benefit and altruism to concerns about the risks of the study.

#### Information, transparency, and accountability for risks

Participants highlighted detailed information and transparent communication as crucial factors influencing their participation in research. In particular, they called for clear explanations of potential risks and benefits, as well as reliable mechanisms for protection and recourse in the case of unanticipated harms. One participant said, “If you gave us the explanation, the paper here [informed consent form], which we will be signing and reading, and it said there that you would be responsible for anything serious or any mistake that happened to us, then yes, I would participate.” Another participant described the importance of dialogue and engagement with PDL to achieve consensus before conducting vaccine trials in prisons: “The vaccine could start inside the prisons, (…) if they came here, brought everyone together and reached a consensus. It’s not just about coming here without giving any explanation, simply saying, let’s vaccinate everyone for tuberculosis”.

#### Right to decide to participate

A central theme in the focus groups was the declaration of one’s right to make decisions about participating in research. One participant advocated for autonomy in making decisions that would affect her own health: “Whether I participate in this study is my decision…even though I’m incarcerated. Because it’s my health, right?” Another participant claimed the right to participate in research as a human right to which she, “like any citizen,” was entitled, despite being deprived of the right to liberty (“to return to our home”). This sentiment was corroborated by yet another participant who alluded to a scenario wherein incarcerated individuals and community members alike were eligible to participate in vaccine trials: “Just as it’s doing us good, it will also be doing our family good…I take it [the vaccine] here, she takes it [the vaccine] there.” Such attitudes were prevalent throughout the focus groups, revealing a strong consensus around PDL’s perceived ability and right to make informed decisions around participation in research.

#### Respect for Autonomy and Its Challenges in Prisons

Participants reported positive experiences participating in the focus group, emphasizing that their freedom of choice was respected. As one participant expressed, “I think everyone felt comfortable here, because from the beginning the conversation started in the right way… they gave us the chance, do we want to stay or not?” This approach was perceived as honoring each individual’s autonomy, fostering an atmosphere of trust and respect.

At the same time, other accounts revealed that autonomy may be compromised under certain conditions. While participation is often presented as voluntary, some individuals described feeling indirect pressure or fear of consequences for refusing—such as threats to visitation rights. One participant, referring specifically to the context of the COVID-19 pandemic and the vaccination campaign within the prison, stated: “It’s mandatory to take it. Even if someone refuses, if they insist on not taking it, they might lose, lose their visitation rights — as we’ve already been threatened with losing visits. […] That’s why it becomes an obligation.”

### Recommendations for ethical inclusion of PDL in clinical trials of new TB vaccines

Drawing from the perspectives gathered in the FG, we propose a set of stakeholder-informed recommendations to guide the inclusion of PDL in clinical trials for new TB vaccines.

These recommendations are not intended to be an exhaustive list of necessary conditions; rather, they represent responses to key points raised by PDL, whose voices and perspectives are too often excluded from the research process. The proposed actions include:

1. Provide equitable access to health care irrespective of trial participation.

Ensure universal access to quality health care in prison before, during, and after the trial, for all PDL regardless of trial participation. Substandard care may introduce coercive incentives to participate in research as a means to access better care. Vaccine trials must not exploit or exacerbate existing inequalities in the prison health system.

2. Strengthen education about TB and vaccines to facilitate voluntary, informed consent

Develop consent processes tailored to the prison context, ensuring clarity, autonomy, and the absence of coercion. Information must be accessible, culturally appropriate, and delivered in plain language. Provide additional, independent educational resources about TB, vaccines, and clinical trials to all PDL to enable informed, conscious deliberation around the decision to participate. Highlight that participation is voluntary and that one’s decision to participate will not affect other rights or privileges in the prison.

3. Enact parallel efforts to mitigate TB vulnerability

Evaluate how the heightened vulnerability to TB and fears of infection among PDL may influence their capacity to genuinely weigh risks and benefits when deciding to participate. Incorporate additional safeguards to prevent exploitation of these vulnerabilities, including implementing parallel, prison-wide measures to reduce TB transmission.

4. Actively involvePDL in study design and oversight

Incorporate the voices and lived experiences of PDL in all phases of the study, especially the design and implementation phases. Structured mechanisms such as community advisory boards, as well as more informal processes like open forums or discussion circles, serve to promote transparency, trust, and accountability. During the active phase of the study, these mechanisms also provide an ongoing channel of communication to inform stakeholders about study progress and results, proactively address concerns, and seek feedback and input for protocol modifications if necessary.

5. Guarantee post-trial access to effective vaccines

If a vaccine candidate is proven safe and effective, all participants must be granted priority access to the new TB vaccine once approved. Detailed vaccine allocation policies and logistical frameworks must be established in advance to ensure timely access for PDL to a successful vaccine following trial completion.

6. Recognize the autonomy and right of PDL to participate in science

Respect the right of PDL to make informed decisions about their own health and to access scientific advancements, including through participation in research. While imprisonment may entail the loss of certain rights, it should not automatically preclude PDL from exercising their autonomy in deciding whether to participate in research. Ethical challenges surrounding the inclusion of PDL in clinical trials–rather than serving as blanket justifications for exclusion–should be met with comprehensive measures, strong safeguards, and effective oversight mechanisms developed in partnership with PDL and their advocates.

## Discussion

In ethical debates around the inclusion of PDL in vaccine trials for TB and other diseases, it is crucial to center the perspectives of those with lived experience of incarceration. However, these perspectives remain poorly understood, particularly in low- and middle-income countries such as Brazil. This qualitative study aimed to understand the perceptions and experiences of PDL across Brazil regarding TB, new TB vaccines, and their willingness to participate in vaccine trials. Participants articulated a strong desire to protect themselves, their families, and their communities from TB, recognizing both the severity of the disease and the significance of research on new vaccines against TB. Despite the challenges inherent to carceral environments, many expressed a willingness to participate in TB vaccine trials, conditional upon meaningful engagement of PDL, clear communication and transparency, and strong safeguards to enable autonomous and coercion-free participation. Willingness to participate in TB vaccine trials was high among FG participants, driven by a combination of altruism, personal and familial protection, and a desire to contribute to broader public health efforts.

Notably, participants defended their ability to make conscious decisions around participation in research and declared their right to access participation in science. They pointed to their experience with the present study as an example of research that respected their autonomy through transparency, clear communication, and the freedom to decline participation. They also highlighted the importance of active engagement of PDL throughout the research process and the need for clear mechanisms for protection and recourse in case of harm. These findings suggest that, despite the inherent vulnerabilities associated with incarceration, PDL view their inclusion in research as ethically possible and a right to which they are entitled, provided that rigorous safeguards are upheld.

A complicating factor is the current state of prison healthcare in Brazilian prisons. Participants reported difficulties accessing essential care and medicines, indifference and neglect from prison staff, a lack of reliable information, and significant fears of TB. Similar findings of inadequate healthcare services and systemic neglect have been reported in surveys from prisons throughout the region (8,30). These findings underscore the necessity of improving healthcare access for all PDL to prevent coercive influences on research participation and to ensure quality care before, during, and after a research study.

Relatedly, we found pervasive fears of TB and limited access to TB information and services, which may generate pressure to participate in trials due to the absence of alternative preventive measures. To mitigate this potential source of coercion, it is essential to implement parallel efforts to reduce the exorbitant TB risk in prisons. Furthermore, the lack of accurate and accessible information about TB must be addressed to enable PDL to fully and appropriately weigh the risks and benefits of trial participation. Educational initiatives led by trusted health professionals—identified in other prison-based studies as critical for fostering informed decision-making and building trust—are likely to be essential(31–33).

While incarceration entails certain restrictions, the right to health—enshrined in international human rights frameworks—must be fully upheld(19). This includes not only access to TB care and services but also the right to participate in scientific research and benefit from its advances. The United Nations Mandela Rules affirm that persons deprived of liberty retain all fundamental rights, including access to health and scientific progress, consistent with human dignity(34). The right to science, as outlined in international covenants such as the ICESCR (Article 15), includes both the right to benefit from scientific advancement and to participate in its development (35).

To date, however, participation in research has been severely restricted for PDL due to carceral conditions that impede voluntary, informed, and ethically sound engagement. As discussed in our forthcoming *Lancet Infectious Diseases* perspective, this exclusion reinforces systemic inequities in who has access to medical advancements and contributes to the invisibility of vulnerable populations in scientific agendas. These barriers, though real, must not be used as justification for the continued systematic exclusion of incarcerated individuals from science. Instead, they underscore the need to build frameworks that enable ethical inclusion with transparency, autonomy, and protection.The recommendations outlined in this study, informed by the perspectives and attitudes of PDL, provide guidance for future strategies to enable the ethical inclusion of PDL in TB vaccine trials, respecting their autonomy and upholding their right to participate in scientific research (35).

This study has several limitations. Convenience sampling may have limited the representativeness of our findings, although we aimed to include PDL with varying demographics and from different housing units. Our results may nonetheless have limited generalizability to settings with different TB epidemiology, prison conditions, and cultural contexts. Future studies may explore these topics in surveys with larger, more heterogeneous samples and settings.

Additionally, our findings may be subject to social desirability bias, as participants might have felt compelled to offer responses that aligned with perceived expectations of facilitators or peers. The group setting may also have constrained open discussion on sensitive topics, including potential experiences of coercion, distrust, or pressure from prison authorities, gang dynamics, or other PDL. In highly stratified prison environments, group hierarchies and social tensions may influence the expression of dissent, fear, or more critical perspectives on participation in vaccine trials. Future studies could consider using complementary methodologies—such as anonymous surveys or in-depth individual interviews—to elicit more candid responses and triangulate findings across formats.

## Conclusion

This study investigated the perceptions of PDL in Brazil regarding research on TB and vaccines, aiming to understand their knowledge, experiences, and challenges in this context. Our qualitative analysis revealed several potential sources of structural coercion for vaccine trials in prisons, including unmet health needs, prevalent fears of TB, neglect by prison staff, and limited access to information. Nonetheless, many individuals expressed interest in participating in TB vaccine trials, conditional upon clear, transparent information, engagement of PDL, protection from coercion, and recourse in case of harm. Comprehensive efforts to fulfill these conditions are urgently needed to enable the ethical inclusion of PDL in clinical trials and to promote their equitable access to scientific innovations.

## Data Availability

The data that support the findings of this study are available from the corresponding author upon reasonable request, in accordance with ethical guidelines and participant confidentiality.

## Funding

This work was supported through Open Philantropy.

## Acknowledgements

The authors thank the people deprived of liberty who agreed to participate in the study, to the state health and public security departments and research centers for their full support during the study period. We would also like to thank the funding body (Open Philanthropy).

## Authors’ contributions

Conceptualization: J.R.A., C.C.M.G., and J.C.; methodology: E.F.L., L.F.S., J.R.A., C.C.M.G., and J.C.; validation: L.F.S., J.R.A., C.C.M.G., and J.C.; formal analysis: M.C.C.F.P., Y.E.L., E.F.L., L.F.S., M.M., M.G.C., J.R.A., C.C.M.G., and J.C.; investigation: M.C.C.F.P., Y.E.L., E.F.L., L.F.S., M.M., M.G.C., J.R.A., C.C.M.G., J.C., D.B.P., M.P.A.V., R.R., S.M.B., G.L.F., M.C.S., J.S.P., G.P.S-M., L.B.O.A., L.G.P.,T.A.H., D.K.B., and P.S.; resources: J.R.A., E,FL., C.C.M.G., and J.C.; data curation: M.C.C.F.P., Y.E.L., E.F.L., L.F.S., and M.G.C.; writing—original draft preparation: M.C.C.F.P., Y.E.L., E.F.L., L.F.S., M.M., M.G.C., J.R.A., C.C.M.G., J.C., D.B.P., G.L.F., M.C.S., and L.G.P.; writing—review and editing: M.C.C.F.P., Y.E.L., E.F.L., L.F.S., M.M., M.G.C., J.R.A., C.C.M.G., J.C., D.B.P., M.P.A.V., R.R., S.M.B., G.L.F., M.C.S., J.S.P., G.P.S-M., L.B.O.A., L.G.P.,T.A.H., D.K.B., and P.S.; visualization: M.C.C.F.P., Y.E.L., E.F.L., L.F.S., M.M., M.G.C., J.R.A., C.C.M.G., J.C., D.B.P., M.P.A.V., R.R., S.M.B., G.L.F., M.C.S., J.S.P., G.P.S-M., L.B.O.A., L.G.P.,T.A.H., D.K.B., and P.S.; supervision: M.C.C.F.P., Y.E.L., E.F.L., L.F.S., M.M., M.G.C., J.R.A., C.C.M.G., J.C., D.B.P., M.P.A.V., R.R., S.M.B., G.L.F., M.C.S., J.S.P., G.P.S-M., L.B.O.A., L.G.P.,T.A.H., D.K.B., and P.S.; funding acquisition:J. R. A., J.C. All authors have read and agreed to the published version of the manuscript.

## Conflicts of interests

The authors declare that they have no competing interests.

## Notes

### Competing Interest Statement

The authors have declared no competing interest.

### Author Declarations

This study was approved by the following institutional review boards and ethics committees: Ethics Committee for Research (CEP) of Federal University of Mato Grosso do Sul – UFMS (Approval No. 6.612.127, CAAE: 75865823.7.1001.0021, January 12, 2024) Center for Research in Tropical Medicine of Rondônia – CEPEM (Approval No. 6.633.785, CAAE: 75865823.7.2001.0011, February 3, 2024) School of Nursing at the Federal University of Minas Gerais – UFMG (Approval No. 6.708.897, CAAE: 75865823.7.2002.5149, March 18, 2024) University of Santa Cruz do Sul – UNISC (Approval No. 6.711.854, CAAE: 75865823.7.2005.5343, March 19, 2024) Tropical Medicine Foundation Doctor Heitor Vieira Dourado – FMT-HVD (Approval No. 6.745.992, CAAE: 75865823.7.2004.0005, April 5, 2024) All participants provided voluntary written informed consent prior to participation. The study also received authorization from the respective State Departments of Penitentiary Administration.

